# Insufficient social distancing may be related to COVID-19 outbreak: the case of Ijuí city in Brazil

**DOI:** 10.1101/2020.06.22.20132910

**Authors:** Thiago Gomes Heck, Rafael Zancan Frantz, Matias Nunes Frizzo, Carlos Henrique Ramires François, Mirna Stela Ludwig, Marilia Arndt Mesenburg, Giovano Pereira Buratti, Lígia Beatriz Bento Franz, Evelise Moraes Berlezi

**Affiliations:** Research Group in Physiology, Department of Life Sciences, Regional University of Northwestern Rio Grande do Sul State (UNIJUÍ), Ijuí, RS, Brazil; Postgraduate Program in Integral Attention to Health (PPGAIS-UNIJUÍ/UNICRUZ), Ijuí, RS, Brazil; Medicine Course, Department of Life Sciences, Regional University of Northwestern Rio Grande do Sul State (UNIJUÍ), Ijuí, RS, Brazil; Department of Exact Sciences and Engineering, Regional University of Northwestern Rio Grande do Sul State (UNIJUÍ), Ijuí, RS, Brazil; Federal University of Pelotas (UFPel) and Federal University of Health Sciences of Porto Alegre (UFCSPA); Research Group in Human Aging, Department of Life Sciences, Regional University of Northwestern Rio Grande do Sul State (UNIJUÍ), Ijuí, RS, Brazil

**Keywords:** COVID19, social distancing, coronavirus, mobile monitoring

## Abstract

The coronavirus disease initiated in 2019 (COVID-19) has proven to be highly contagious and quickly became a pandemic. Nowadays, it presents higher transmission rates worldwide, chiefly in small Brazilian cities, as Ijuí. Located in the northwestern of the State of Rio Grande do Sul (RS) with 83,475 inhabitants, Ijuí was selected to receive a population-based survey organized in five steps, involving 2,222 subjects. Subjects were tested for the presence of antibodies against coronavirus (SARS-CoV-2) and answered questions regarding social distance adherence (SDA), daily preventive routine (DPR), comorbidities, and sociodemographic characteristics. In parallel, the local government registered the official COVID-19 cases in Ijuí, and the mobile social distancing index (MSDI) was also registered. In this study, we demonstrate the decrease in the levels of SDA, DPR and MSDI before the beginning of COVID-19 community transmission in Ijuí. Also, we provide predictions for cases, hospitalization, and deaths in the city. We concluded that the insufficient social distancing, evidenced by different methods, might have a strong relationship with the rapid increase of COVID-19 cases in Ijuí. Our study predicts a closer outbreak of community infection of COVID-19, which could be avoided or attenuated if the levels of the social distancing in the population increase.

## Introduction

At the end of 2019, some cases of pneumonia of “unknown cause” were noticed by the Wuhan Municipal Health Commission (China) [1]. Collected bronchoalveolar-lavage samples were consistent with an RNA virus of the Coronaviridae family [2]. Thereafter, The World Health Organization named the novel infectious pneumonia as “coronavirus disease 2019” or COVID-19 [3]. COVID-19 had proven to be highly contagious by the time it reached approximately 300 cases in China in January 2020 [4]. Thus, it became an epidemic case, with more than 44,000 infections and more than 1,000 deaths in China, with 441 cases outside China in 24 countries [5].

The first case of COVID-19 in Brazil was reported on February 27^th^ in the city of São Paulo. Based on published events, eight of the twenty-seven National Federation Units present cumulative mortality rates above 10 per 100,000 inhabitants: four in the North, two in the Northeast, and two in the Southeast Region (Rio de Janeiro and São Paulo) [6]. In the state of Rio Grande do Sul (RS), the southernmost State in Brazil (with 11.3 million people), the first case of COVID-19 was diagnosed on February 29^th^ 2020. As of August 6^th^, 76,563 confirmed cases (673 per 100,000 inhabitants), and 2,163 deaths (19 per 100,000 inhabitants, 2.8% of confirmed cases) had been reported [7,8].

Therefore, in the RS, a population-based survey in nine sentinel cities (EPICOVID-RS) was conducted in a period from April to June [8] and included the city of Ijuí, which is localized in the Northwestern region of the state, with a population of 83,475 inhabitants. In this survey, we tested 2,500 subjects for the presence of antibodies against SARS-CoV-2 divided into five rounds (5 with ~500 subjects each), indicating that the seroprevalence was undetectable (0 positive cases) on April 12^th^ (round 1), and increased to 0,042%[7,8]. Questions regarding social distancing adherence (SDA) and daily preventive routine (DPR) were answered by the subjects who participated in the survey [8]. In parallel, the local government registers and turns public the official COVID-19 cases in Ijuí (for details, see the official government website [9]), while the mobile social distancing index (MSDI) in Ijuí was monitored by means of data collected from mobile geolocation [10].

Since there are still no pharmaceutical treatments available, interventions have focused on quarantine and social distancing. The aim of these strategies is to slow down the spread of infection and reduce the intensity of the transmission (a.k.a “flatten the curve”)[11]. The Wuhan outbreak indicates that critical care capacities can be exceeded many times if distancing measures are not implemented quickly or strongly enough [12]. An effective social distancing strategy and monitoring may reduce the risk of an overwhelming the health system, allowing adequate patient care and decreasing mortality rates [12]. Thus, considering the local COVID-19 data and the social distancing behavior registered, we showed that low levels of social distancing were not enough to avoid the beginning of the COVID-19 outbreak in Ijuí.

## Materials and Methods

### Subjects, ethics and study design

Ijuí (latitude 28°23'16 to the south and a longitude 53°54'53" west) is the most populous city in the Northwest region of the state. It is considered a city of students (“University city” with 83,475 residents) and a center of hospital and University resources, as well as the largest and most important population center in the region, with a population rounding 150,000 people.

In this study, we follow an interdisciplinary approach to investigate whether the behavior of citizens is associated with the increase in the number of COVID-19 outbreak in Ijuí. First, we considered official data about COVID-19 cases, which is daily updated by the public administration on the government website and plotted as a cumulative case line graph by the date of the initial symptoms and the date of the confirmed test [9]. Second, we analyzed the MSDI for a period that comprehends February 1^st^ to July 5^th^ 2020 [10]. Third, we analyzed data from the study EPICOVID-RS in Ijuí regarding SDA and DPR [8]. Finally, we make predictions (starting at 100^th^ case) for COVID-19 in Ijuí regarding the number of cases, hospitalizations, and deaths, considering different scenarios of transmission rates and compare to update COVID-19 data.

The state of RS is divided by the National Institute of Geography and Statistics [13] in eight intermediary regions, so the main city in each region was selected for the study EPICOVID-RS [8]. Ijuí is one of the main cities, so it was selected for this study. Ethical approval was obtained from the Brazilian National Ethics Committee (process number 30415520.2.0000.5313), with written informed consent from all participants.

### Official COVID-19 data in Ijuí

The COVID-19 Municipal Scientific Committee of Ijuí developed an information panel, with public data from the Municipal Epidemiological Surveillance. With this statistical data, the population receives all the information regarding the number of cases confirmed, hospitalizations, recovered, and deaths [9]. The number of cases confirmed is also discriminated by age and gender. Confirmed cases consist of data received from the laboratory analysis, including RT-PCR, Rapid Tests, and Chemiluminescence tests. The numbers of confirmed cases published in the bulletins of the municipal health department may differ from those reported by the state information system (e-SUS VE), because of the frequency of updating the system. Furthermore, the number of confirmed cases shown by the reports of Ijuí Hospitals may differ from municipal data because of the origin of hospitalized patients, which are form different neighboring cities. Our study takes into consideration only data for people residing in Ijuí and from the Ijuí Municipal Health Department.

### Mobile Social Distancing Index (MSDI)

The MSDI was created by the Brazilian private company Inloco (www.inloco.com.br), which has developed a technology to register fingerprints based on geolocalization, which creates a unique identity to users of mobile devices [10]. The company currently monitors a stable set of 60 million mobile devices spread all over the cities of the country. Every device stores and sends anonymous location data regarding its movement during the day every time it connects to the Internet. The MSDI is computed on a daily basis and comprehends the percentage of mobile devices that in a given municipality remain within a radius of 450 meters from the point identified as home, with a precision of the system being less than three meters [10]. Within the period considered in this study (from February 1^st^ to July 5^th^ 2020), the dataset has a total of approximately 1 million records for every city in Brazil. We extracted from this dataset the MSDI the city of Ijuí grouped in a daily measurement.

### Population-based survey protocol: EPICOVID-RS in Ijuí

Multistage sampling was used to select census tracts with probability proportionate to size in the city, and ten households at random in each tract based on census listings updated in 2019. All household members were listed at the beginning of the visit, and one individual was randomly selected through an app used for data collection. The survey rounds took place in the following periods: April 11-13^th^ and 25-27^th^, May 9-11^th^ and 23-25^th^, and June 27-29^th^ (for data presentation in figures we considered the mean value for the three days interval). If there were any refusals at the household level, volunteers were instructed to select the next household on the list until they have visited ten families. In the 2^nd^ wave, field workers went to the house visited in the first wave, and then selected the tenth household to its right. The same procedure was performed in the last three waves. In case of refusal, the next house to the right side was selected. In the case of acceptance at the household level, but the selected individual refused to provide a sample, a second member was selected. If this person also refused, the field workers moved on to the next household on the list. (for more details, see reference [8]). Prevalence of antibodies was assessed with a rapid test using finger-prick blood samples - the WONDFO SARS-CoV-2 Antibody Test (Wondfo Biotech Co., Guangzhou, China). The sensitivity and specificity of this rapid test were previously validated [8,14].

Participants answered short questionnaires, including sociodemographic information (sex, age, diseases, schooling, and skin color), COVID-19-related symptoms, use of health services, compliance with social distancing measures, and use of face masks. Field workers used tablets or smartphones to make a full audio record of each interview, as well as, to register all answers and photograph the test results. All positive or inconclusive tests were read by a second observer, as well as 20% of the negative tests. If the selected subject in a household had a positive result, all other family members were invited to be tested. In this study, we described the characteristics of the sample regarding sex, age, education, ethnicity, and comorbidities (mean and upper and lower limit of confidence interval). The focus of our analyses was on declared social distancing behavior and daily behavior characteristics, as well as the association between comorbidities and social distancing behavior using logistic regression and describing as odds ratio and upper and lower limit of the confidence interval. Interviewers were tested and found to be negative for the virus and so provided with personal protection equipment that was discarded after visiting each home. Positive cases were reported to the municipal and statewide COVID-19 surveillance system. The number of COVID-19 positive subjects was noticed in social media by the RS government always three days after each wave.

### Prediction of COVID-19 cases, hospitalizations, and deaths

Estimative of new cases were first performed using real data about COVID-19 from March 22^th^ to May 17^th^ (nine points). Predictions were calculated using equations generated as follows: exponential, defined as proportional rate growth using the expression y = *Y*0 – (*V*0/*K).(1 - e^-kx^*). Alternatively, linear, defined as a straight line using the expression *y = mx + c*. After curve fitting, correlations were predicted (extrapolated) by using the online version of MyCurveFit software (https://mycurvefit.com/), and the results were plotted using GraphPad 8.0.

To estimate the number of exposed subjects, COVID-19 cases, hospitalization rates, and deaths after the 100^th^ case of COVID-19 in Ijuí, we used the method SEIR (Susceptible → Exposed → Infected → Recovered) model [15]. The full description of the equation was provided in the manuscript of WU and Colleagues [16]. We used the following parameters for the predictions: population 83,200; an initial number of cases as 100; *R*_0_ = 2.79 as the basic reproductive number that represents the number of secondary infections that each subject produce, based on the transmission rate documented by the time of the study as a median of 12 studies [17,18]. Also, we used the reproduction number estimated for RS (*R*_0_ = 1.44) on May 8^th^ [19].; incubation of virus time as 5.21 days; time that patient remains transmitting virus infection as 2.3 days; the time between incubation and death of 30 days; hospitalization days until recovery as 21 days (severe) and 14 days (mild); hospitalization rate as 12% and mortality at 2.3%, based on RS governmental data until the end of June [7]; time of hospitalization as 14 days. We used these parameters to calculate the number exposed subjects, COVID-19 cases, hospitalization rates and deaths after the 100^th^ case of COVID-19 in Ijuí in six different situations: 1) maintaining the transmission rate of community infection by COVID-19 as *R*_0_ = 2.79; 2) reducing by 50% the transmission rate (*R*_t_ = 1.44) of COVID-19 exactly 30 days after the 100^th^ case; and, 3) reducing by 50% the transmission rate (*R*_t_ = 1.44) of COVID-19 exactly 15 days after the 100^th^ case. 4) maintaining the transmission rate of community infection by COVID-19 as *R*_0_ = 1.44; 5) reducing by 50% the transmission rate (*R*_t_ = 0.79) of COVID-19 exactly 30 days after the 100^th^ case; and, 6) reducing by 50% the transmission rate (*R*_t_ = 0.79) of COVID-19 exactly 15 days after the 100^th^ case. These estimations were calculated using a free website of the University of Sao Paulo for epidemic profile calculation [20].

## Results

The daily MSDI computed using data from mobile monitoring for the city of Ijuí is showed in Figure 1A. The period of monitoring revealed that on the first day of mobile monitoring, the MSDI was 30.7% (Saturday) and reached 39.3% on Sunday (February 2^nd^) and decreased to 27.5% in the first working day registered (February 3^rd^). These data describe the normal social behavior in Ijuí, which remained with few fluctuations in average until mid-March, when it scales up to 70.2% on Sunday, (March 22^th^) and sustained between 50% and 55% on subsequently working days. However, at the end of June, the MSDI decreased to 31.4% (on June 19^th^), reaching the lowest value since the first case in the city (registered as March 18^th^, see Figure 1B). The last register in working data showed 35.3% of MSDI. For a fair comparison, percentages pointed above the open circles on Figure 1A correspond to MSDI on weekends, whereas the closed circles correspond to MSDI on working days. Thus, it is possible to observe that there is naturally an increase in distancing due to the fact that more people tend to stay in their homes on weekends. However, the decrease in MSDI may be observed, including on weekends since was registered a 70.2% on March 22^nd^, while this level decreased to 48.2% on June 21^st^. From a macro point of view, it is evident that the population of Ijuí is leaving distancing and increasing its social interaction up to the end of June.

The first COVID-19 case in Ijuí (Figure 1B) started to feel symptomatic on March 18^th^ (diagnosed after that). It is possible to observe that until May 17^th^, there was a slight increase in the number of cases registered in Ijuí (Figure 1B). However, on June 7^th^, there was a 2.1-fold increase in total COVID-19 cases. The profile of the confirmed cases revealed that 52.4% of the COVID-19 cases in Ijuí occurred in women and 47.7% in men, and the majority of cases (86%) are younger than 60 years old (Figure 1C). Until the end of June, Ijuí presented a 9,5% hospitalization of positive COVID-19 cases, which 74.1% are older than 50 years, and there was no hospitalization in subjects younger than 30 years old. The majority (70.4%, n = 19) of the hospitalized cases are men in comparison with women (29.6%, n = 8). Also, until the end of June, there was just one death caused by COVID-19 in Ijuí (men, 85 years-old, oncologic patient with diabetes and hypertension). The urban distribution of COVID-19 cases in Ijuí indicates that approximately 30% are downtown inhabitants (Supplementary material, Figure S1).

**Figure 1:**
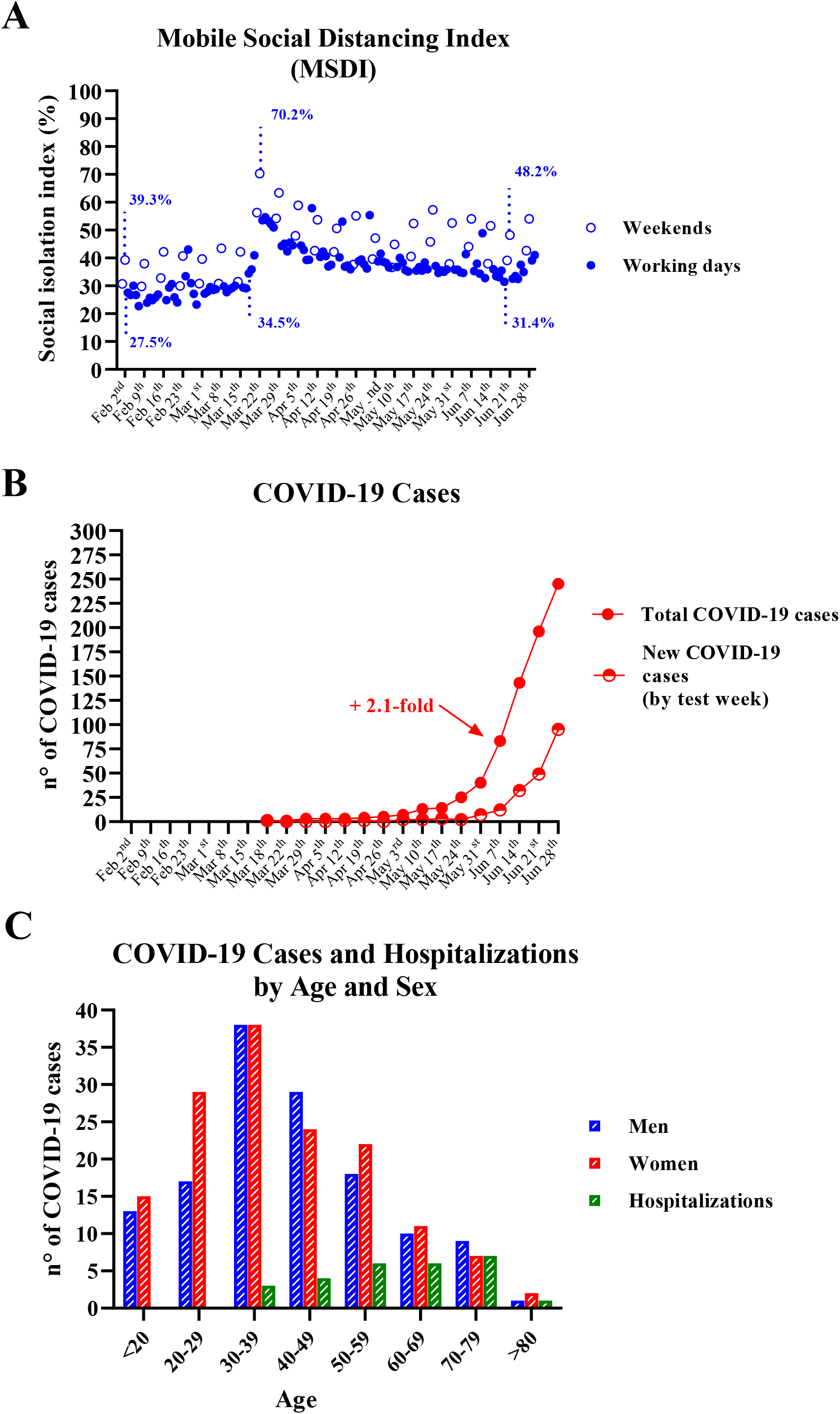
Mobile social distancing index and COVID-19 cases in Ijuí-Brazil. **A)** Mobile Social distancing index (MSDI) **B)** Total and new weekly COVID-19 cases **C)** Distribution of COVID-19 cases and hospitalizations by age.

In each round of the population-based survey, more than 400 adults were inquired and tested to the presence of Sars-Cov-2 antibodies, totalizing 2,222 subjects included in the study. Characteristics of the subjects are described in Supplementary material Table 1 and Table 2 as supplementary material. The social distancing population-based survey started on April 12^th^, and revealed that the majority of the subjects declared high SDA to the recommendations (Figure 2A). However, it is possible to observe a decrease in the proportion of subjects that declared adherent to social distancing (high adherence) in parallel to the increase of the proportion that partially adheres to social distancing (Figure 2A). Similarly, the proportion of the subjects that stayed at home during all the time or declared go out only for essential needs has decreased, while there was an increase in the number of people that declared went out their home daily for work (Figure 2B). Thus, it is possible to observe that there was a decrease in the MSDI just 15 days after the first round, which remains to decrease until the end of May (blue lines in Figure 2C). In contrast, there was a significant increase in the number of COVID-19 cases in Ijuí in the period of May to June (red line, Figure 2C), which may evocate a recovered of SDA and DPR approximately one month later (June) (blue lines in Figure 2C).

**Figure 2.**
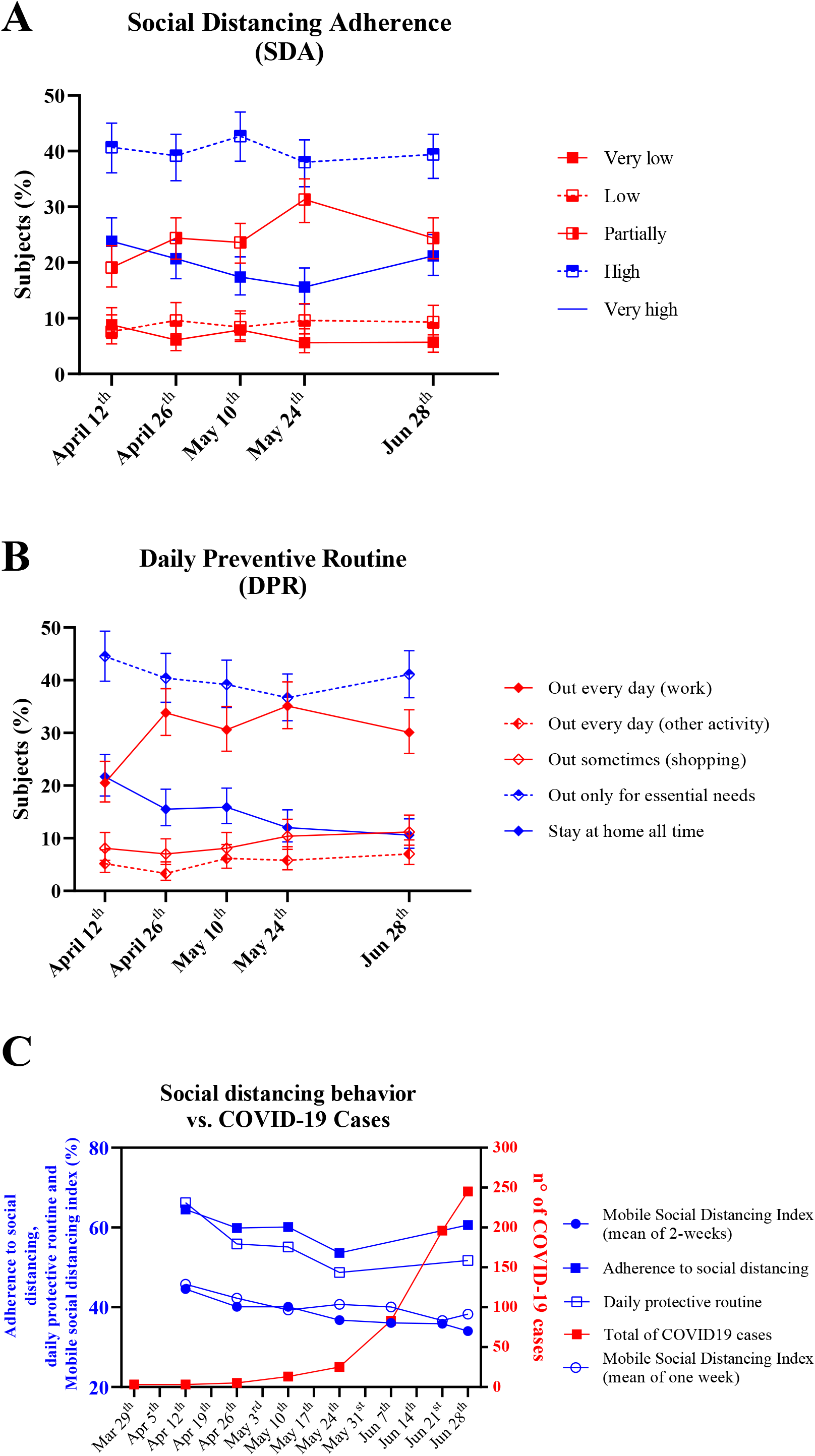
Social distancing adherence, daily preventive routine, and mobile social isolation index association with COVID-19 cases in Ijuí-Brazil. **A)** Social distancing adherence indicated in the population-based survey study. **B)** Daily preventive routine indicated in the population-based survey study **C)** Association between total COVID-19 and social distancing data.

We have also analyzed two scenarios to estimate the number of cases in Ijuí after May 17^th^ (Figure 3A): First, a sustained controlled community infection represented by a linear trend (green line in Figure 3A, estimated by the equation y = 1.416667x −1.416667, R² = 0.848, P = 0.0004); second, an uncontrolled outbreak community infection in Ijuí, represented by an exponential progression in the number of cases (blue line in Figure 3B, y = 1.351649 - (−0.2424262/-0.3155474).(1 - ^e^(+0.3155474x)^, R^2^ = 0.955, P = 0.00019). Worryingly, the number of real cases registered (red line in Figure 3A) was higher than estimated by linear and exponential prediction, since 277 COVID-19 cases registered in Ijuí until July 7^th^, while the estimative predicted 23 and 120 COVID-19 cases by linear and exponential equations, respectively. Thus, since the exponential estimative (blue line) showed that Ijuí could reach 1,000 cases on August 23^rd^, the anticipation of this scenario, marked by the real registered COVID-19 cases, indicates a risk to overwhelm the public health system in Ijuí.

Considering the real scenario in the context of the 100^th^ case of COVID-19 in Ijuí, we analyzed the predictions for the following months after that. It is possible to estimate a great effect on the number of subjects exposed to coronavirus in the next two months, with higher levels of infected subjects, hospitalization, and deaths, if the transmission rate is maintained, while would have a great benefit if decreases the transmission rate as soon as possible, mainly if reach controllable levels. In this worst scenario (*R_0_* = 2.79), the overwhelm of public and private health care systems is evident since it would more than 4,000 hospitalizations cases simultaneously and more than 1,500 deaths in the following 120 days (Figure 3B). In contrast, if the transmission rate of COVID-19 is reduced by 50%, precisely 30 days after the 100^th^ case (*R_t_* = 1.40), it is expected a moderate decrease in the impact on the healthcare system (Figure 3C). For further protection, we analyzed the impact on the transmission rate of COVID-19 if the population reduces by 50% (*R_t_* = 1.40), precisely 15 days after the 100^th^ case (Figure 3D). Thus, the exposed subjects would decrease, reducing the peak of infected subjects to 1,314, expected to occur in 66 days. Consequently, the peak of hospitalization would be later, close to three months after the 100^th^ case (85^th^ day), with 1,582 subjects, resulting in 862 deaths in 120 days. If we consider the RS transmission rate document (*R_0_* = 1.44) without any alteration after the 100^th^ case would be found a higher number of infections, hospitalizations, and deaths peaking after two months (Figure 3E). However, if the transmission rate of COVID-19 is reduced by 50%, precisely 30 days after the 100^th^ case (*R_t_* = 0.79), it is expected a considerable decrease in the impact on the healthcare system, with 156 daily cases in the peak (34^th^ day) and 120 hospitalizations by day two months after the 100^th^ case, and 62 deaths in 120 days (Figure 3F). Finally, a significant decrease would be expected if the transmission rate of COVID-19 reduces by 50% (*R_t_* = 0.79) precisely 15 days after the 100^th^ case (Figure 3G): the infection peak would reach 75 simultaneous cases and 52 hospitalizations, resulting in 27 deaths. The proportion of the whole population that needs recovered decreased from approximately 82.0% in the worst prediction to 1.37% in the best one. In detail, in the last prediction, considering a reduction of 50% in the transmission rates, if applied 15 days after the 100^th^ case, approximately 98.7% of the population of Ijuí would remain without infection, and approximately 1.37% would be recovered (1,162 subjects).

**Figure 3.**
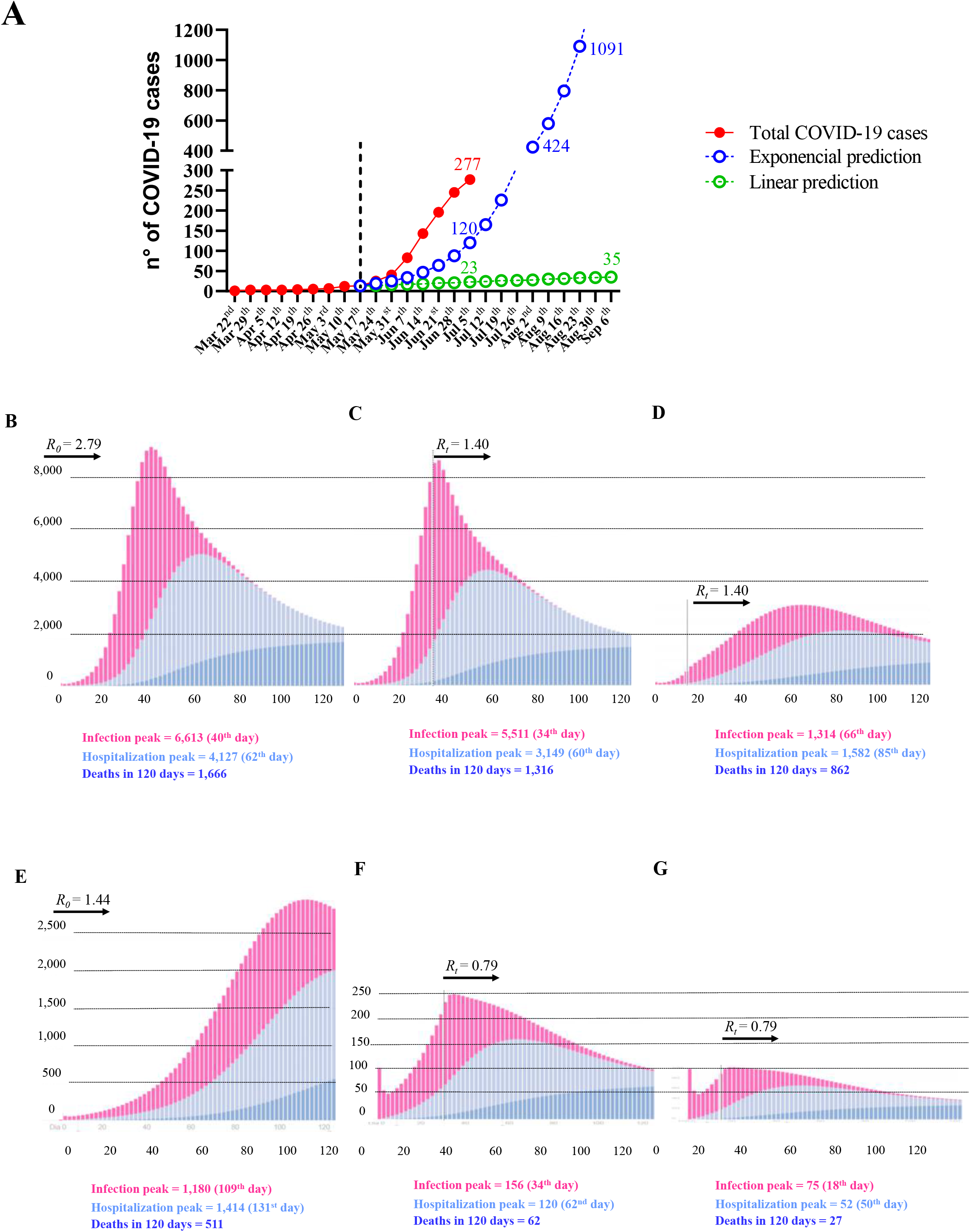
Predictions for COVID-19 cases, hospitalizations, and deaths in Ijuí-Brazil. **A)** Difference between real cases and predictions of COVID-19 using linear regression (green line) and exponential equation (Blue line). **B)** Predictions without any new intervention after the 100^th^ case (*R_0_* = 2.79). **C)** Predictions after implementing strategies that reduce 50% in the transmission rate 30 days after the 100^th^ case (*R_t_* = 1.40). **D)** Predictions after implementing strategies that reduce 50% in the transmission rate 15 days after the 100^th^ case (*R_t_* = 1.40). **E)** Predictions without any new intervention after the 100^th^ case (*R_0_* = 1.44). **F)** Predictions after implementing strategies that reduce 50% in the transmission rate 30 days after the 100^th^ case (*R_t_* = 0.79). **G)** Predictions after implementing strategies that reduce 50% in the transmission rate 15 days after the 100^th^ case (*R_t_* = 0.79). All predictions were performed in the epidemic simulator free app from São Paulo University. Available on https://ciis.fmrp.usp.br/covid19/epcalc/public/index.html

## Discussion

Our study described the SDA, DPR, MSDI, predictions, and the progression of COVID-19 cases in Ijuí (Brazil). To the best of our knowledge, this is the first study that described the social distancing behavior of the community based on a population-survey procedure together mobile monitoring data, that preceded the COVID-19 outbreak. Also, it is the first report regarding estimative of the progression of the disease in the city.

During February, the city of Ijuí, as well as its neighboring cities, have not registered any case of COVID-19 in their population. The first case registered in the estate of RS, to which Ijuí belongs, was in the city of Campo Bom, 402 km away from Ijuí, and very close to the capital of the estate (Porto Alegre), on March 10^th^. The first case of COVID-19 registered in Ijuí dates back to March 18^th^, when the patient started to feel symptomatic. Note that, only after eight days Ijuí joined the official counting of cases, and, most probably, the absence of cases before this date explains the non-change in the percentage of social distancing (respected its natural peaks on Sundays).

On March 19^th^ the city administration issued its most restrict decree in the analyzed period, which restricted public transport, closed stores, suspended classes at public and private schools and university, and established protocols for restaurants and other services for one week [9]. This is evidenced when on March 22^nd^ the MSDI reached its highest value, by 70.2%. After that, other decrees were also issued, but with relaxing restrictions. In parallel, the Federal Government of Brazil has been minimizing this pandemic and, in most cases, stimulating the population to keep their regular routine [21]. Thus, after the highest social distancing already registered, it is possible to observe a continuous reduction of the distancing, to the point that at the end of June, we have a value very close to that recorded at the beginning of the analyzed period, when there were no cases in Ijuí and not even in Brazil [22,23].

In the last days of June, it is possible to observe a slight increase in the distancing, most probably a consequence of a high number of new cases reported in the second half of June, which have demanded the city administration to review its protocols and be stricter [9]. The mean of the MSDI for the total period of mobile monitoring is 38.5%. If we take into consideration that was classified as “stayed at home” when the mobile did not move more than 450m (radius) [10], it is possible that the actual social distancing level in Ijuí most probably rounds below 30% in regular times, since this distance allows neighborhood social interactions, including small markets, which strongly points that the social distancing is currently low in the city.

Our results about SDA and DPR may be related to the official recommendations by local and state governments. The state of RS has implemented since May 10^th^, the “Controlled Social Distancing Model” (CSDM), to avoid the COVID-19 outbreak considering recommendations for economic activities. In color flags, which ranges from a yellow flag (to represent a low risk to infection and high health care capacity) to a black flag (to represent a high risk of transmission rate and high occupation rates in the hospitals and intensive care units). Thus, since May 10^th^, every city is classified according to this model to represent the progression of the COVID-19 outbreak [7]. Ijuí was set as a yellow flag from May 10^th^ to June 8^th^, when it became orange (one level above of risk) because of the rapid increase in the number of cases (95 new cases) and the hospitalization rates in one week (1 to 5 subjects) and then decreased the recovered-to-active ratio cases in Ijuí. Also, it was noticed one death and decreased in the intensive care unit vacancy. As a local initiative to counteract the COVID-19 outbreak, the Ijuí Municipal Decree number 7.107 (of June 16^th^) improved the recommendations to avoid community transmission [9]. In detail, the decree allows the maintenance of regular service in restaurants (between 7 a.m and 23 p.m.) with reduced capacity, while snack bars and coffee shops were allowed to work only in the delivery or drive-thru mode. The non-essential commercial was authorized to operate; however, they can only receive one customer per employee, respecting the limits established in the occupation and operation protocols. It is allowed to operate the facilities of sports clubs exclusively for the physical conditioning of the respective contracted professional athletes, observing the minimum distance of two meters between them. Physical contact or agglomerations being prohibited in any case [9]. Thus, the measured social distancing in Ijuí may be in accordance with theses official recommendations but was insufficient to avoid the rapid increase in the number of COVID-19 cases. (For Ijuí urban geographic details, please see Supplementary material figure S1, with geographic localization of COVID-19 cases in June and August).

Social distancing measures adopted by the population appear effective, mainly when implemented in conjunction with the distancing of cases and quarantining of contacts [19,24]. Our data showed that preventive behavior, related to the SDA recommendation and DPR, did not reach 70% of social distancing in any of the four steps of the survey. We mentioned this goal since it was proposed early that maintaining the capacity of social distancing at a maximum of 76% could avoid the death of 90,000 individuals and the overwhelm of the intensive care units in São Paulo (Brazil)[25]. Nowadays, São Paulo registered more than 20,000 COVID-19 cases and 9,00 deaths [23], suggesting that maintaining and strengthening current social distancing measures, quarantining and isolating cases, is absolutely vital to avoid the collapse of the healthcare systems in the country. Also, it has been suggested that a more restrictive recommendation can be more effective in reducing the number of infected subjects [1,25–28].

At the end of June, Brazil reached more than 1.4 million COVID-19 cases [22,23]. The state of RS provides data about COVID-19, which computes on date June 30^th^ 26,941 COVID-19 cases, representing a mean of incidence in the state of 344 cases per 100,000 inhabitants [7]. From June to July, RS decreased the hospitalization rate from 13% to 11%, while increased the death proportion from 2.3 to 2,5% (nowadays reached 8,5 per 100,000 inhabitants). If we consider the same rates of cases, hospitalization, and deaths, Ijuí could have 287 cases, 31 hospitalizations and 7 deaths. In fact, the number of cases (n = 283) is similar than RS prevalence, however, Ijuí has less hospitalizations (n = 27) and deaths (n = 1), and Ijuí ends the month of June with 80.8% intensive care units occupied, with three COVID-19 patients [7,9].

We compare our “optimistic” prediction (reducing by 50%, *R* = 0.79) for 120 days after the 100^th^ COVID-19 case with the nowadays Ijuí presented COVID-19 profile: the infection peak was predicted with 75 simultaneous cases and with 52 hospitalizations, however, Ijuí has now (August 6^th^ means ~60 days after the 100^th^ COVID-19 case) 156 subjects under recovering and 8 hospitalized patients (the total of hospitalized patients until now was 49 of hospitalizations, which means 9,4% of cases). Also, our predictions showed that 120 days after the 100^th^ COVID-19 case, Ijuí would have a total of 27 deaths. Until now, it was registered five deaths. This comparison allows us to infer that the population followed the recommendations to avoid the transmission of the coronavirus, at least in part, since real cases, hospitalizations and deaths are approximately as predicted by a reduction in the transmission rate below 1.0 (*R* = 0.79).

In our study, we showed the profile of hospitalizations by age and sex, and also informed the profile of the subject that was fatally affected by coronavirus. Thus, our study and up-to-date COVID-19 data preliminary indicates an increased risk for developing a severe form of COVID-19 related to the presence of comorbidities, advanced age, and sex (men). It is remarkable that pre-existing comorbidities are strikingly evident (about half of patients), hypertension being the most prevalent (30 %), followed by diabetes (19 %), coronary heart disease (8 %) and 3% of individuals presenting previous pulmonary condition, such as chronic obstructive lung disease [29,30]. COVID 19 patients bearing previous comorbidities were also among those with highest mortality rates, with adjusted OR of 7.42 (CI95%: 6.33-8.79) for hypertensive patients, 9.03 (CI95%: 7.39-11.35) for diabetic subjects, 12.83 (CI95%: 10.27-15.86) for coronary heart disease, while for chronic obstructive lung disease OR is 7.79 (CI95%: 5.54-10.43) [31]. Also, it was reported that 31 % of cases, 45 % of hospitalizations, 53 % of intensive care unit (ICU) admissions, and 80 % of deaths occurred among subjects aged 65 years or older [32]. In the same direction, the case-fatality rates among individuals aged 80 years or older are approximately 20% [33]. It is noteworthy the discrepancy between males (ca. 60 %) and females at the admission in hospitals. Among non-survivors, this difference is even bigger: ca. 70 % are men [1]. In the Lombardy Region, Italy, these figures are still more impressive: 82 % of the patients admitted to intensive care units (ICU) were older men [32]. The increased risk of this profile (male, older and cardiovascular disease subject) may be related to an impaired immune-metabolism cell stress response. Thus, these subjects cannot resolve virus-induced inflammatory burst physiologically (since there is no medicine) being susceptible to exacerbated forms of inflammation [34], which leads to a fatal “cytokine storm” [35,36]. Thus, it is necessary to observe in future studies the adherence to a protective behavior linked to social distancing associated with the presence of comorbidities [3,37]. The preventive behavior in older subjects and people with chronic diseases in the community may avoid higher levels of hospitalization or high mortality rates.

## Conclusion

The insufficient social distancing registered in the population-based study may be related to the rapid increase of COVID-19 cases in Ijuí. These data predict a closer outbreak of community infection of COVID-19, which could be avoided or attenuated if the levels of the social distancing of population increase in the next weeks.

## Data Availability

The COVID-19 data that support the findings of this study are available in government repository, which is daily updated on the government website and predictions were made accordingly the math-epidemiological web sites. All data of this study are available from the corresponding author upon reasonable request.

https://www.ijui.rs.gov.br/noticias/municipio_disponibiliza_painel_de_informacoes_sobre_casos

https://datastudio.google.com/reporting/4ff82b8a-a9ff-4577-b239-da2e38d24443/page/vBjQB

https://ciis.fmrp.usp.br/covid19/epcalc/public/index.html

## Abbreviation list

SDA: Social Distancing Adherence;
DPR: Daily Preventive Daily;
MSDI: Mobile Social Distancing Index;
RS: State of Rio Grande do Sul

## Acknowledgments

We would like to thank all volunteers who performed the population-based survey. We would like to thanks professor Airam Sausen for her COVID19 math prediction classes. This work was supported by the Regional University of Northwestern Rio Grande do Sul State (UNIJUI), Federal University of Pelotas (UFPEL), and Government of Rio Grande do Sul State, as well as by the Coordination for the Improvement of Higher Education Personnel (CAPES).

## Author Contributions

All authors coordinated the five waves of the population-based study performed in Ijuí integrated with EPICOVID-RS study, collecting data about social distancing (exception, MSL and RZF). TGH, RZF, EMB, and MSL performed the analyses and predictions of the COVID19 case, analyzed the governmental decrees, and wrote the manuscript. MAM produced the sociodemographic and EPICOVID-RS data descriptions. TGH and RZF wrote the manuscript. TGH prepared the figures. All authors were involved in analyzing the results and approved the submitted and published versions.

## References

1. Zhou F, Yu T, Du R, Fan G, Liu Y, Liu Z, et al. Clinical course and risk factors for mortality of adult inpatients with COVID-19 in Wuhan, China: a retrospective cohort study. Lancet. 2020. doi:10.1016/S0140-6736(20)30566-3

2. Wu F, Zhao S, Yu B, Chen YM, Wang W, Song ZG, et al. A new coronavirus associated with human respiratory disease in China. Nature. 2020. doi:10.1038/s41586-020-2008-3

3. The epidemiological characteristics of an outbreak of 2019 novel coronavirus diseases (COVID-19) in China. Zhonghua Liu Xing Bing Xue Za Zhi. 2020. doi:10.3760/cma.j.issn.0254-6450.2020.02.003

4. World Health Organization (WHO). Novel Coronavirus (2019-nCoV) Situation Report – 1 21 January 2020. WHO Bull. 2020.

5. (WHO) WHO. Novel Coronavirus (2019-nCoV) Situation Report – 12 1 February 2020. WHO Bull. 2020.

6. Hallal P, Hartwig F, Horta B, Victora GD, Silveira M, Struchiner C, et al. Remarkable variability in SARS-CoV-2 antibodies across Brazilian regions: nationwide serological household survey in 27 states. medRxiv. 2020. doi:10.1101/2020.05.30.20117531

7. Rio Grande do Sul State Governament. Rio Grande do Sul Coronavírus Panel. 2020 [cited 6 Aug 2020]. Available: http://ti.saude.rs.gov.br/covid19/

8. Silveira MF, Barros AJD, Horta BL, Pellanda LC, Victora GD, Dellagostin OA, et al. Population-based surveys of antibodies against SARS-CoV-2 in Southern Brazil. Nat Med. 2020. doi:10.1038/s41591-020-0992-3

9. Ijuí municipal governament. Ijuí city hall official website. 2020 [cited 6 Aug 2020]. Available: https://www.ijui.rs.gov.br/noticias/municipio_disponibiliza_painel_de_informacoes_sobre_casos

10. Peixoto PS, Marcondes D, Peixoto C, Oliva SM. Modeling future spread of infections via mobile geolocation data and population dynamics. An application to COVID-19 in Brazil. PLoS One. 2020. doi:10.1371/journal.pone.0235732

11. Anderson RM, Heesterbeek H, Klinkenberg D, Hollingsworth TD. How will country-based mitigation measures influence the course of the COVID-19 epidemic? The Lancet. 2020. doi:10.1016/S0140-6736(20)30567-5

12. Li R, Rivers C, Tan Q, Murray MB, Toner E, Lipsitch M. Estimated Demand for US Hospital Inpatient and Intensive Care Unit Beds for Patients With COVID-19 Based on Comparisons With Wuhan and Guangzhou, China. JAMA Netw open. 2020. doi:10.1001/jamanetworkopen.2020.8297

13. IBGE. National Institute of Geography and Statistics. 2019 [cited 6 Aug 2020]. Available: https://cidades.ibge.gov.br/brasil/rs/panorama

14. Pellanda LC, Wendland EM, McBride AJ, Tovo-Rodrigues L, Ferreira MR, Dellagostin OA, et al. Sensitivity and specificity of a rapid test for assessment of exposure to SARS-CoV-2 in a community-based setting in Brazil. medRxiv. 2020. doi:10.1101/2020.05.06.20093476

15. Liu Y, Gayle AA, Wilder-Smith A, Rocklöv J. The reproductive number of COVID-19 is higher compared to SARS coronavirus. J Travel Med. 2020. doi:10.1093/jtm/taaa021

16. Wu JT, Leung K, Leung GM. Nowcasting and forecasting the potential domestic and international spread of the 2019-nCoV outbreak originating in Wuhan, China: a modelling study. Lancet. 2020. doi:10.1016/S0140-6736(20)30260-9

17. Adhikari SP, Meng S, Wu YJ, Mao YP, Ye RX, Wang QZ, et al. Epidemiology, causes, clinical manifestation and diagnosis, prevention and control of coronavirus disease (COVID-19) during the early outbreak period: A scoping review. Infectious Diseases of Poverty. 2020. doi:10.1186/s40249-020-00646-x

18. Riou J, Althaus CL. Pattern of early human-to-human transmission of Wuhan 2019 novel coronavirus (2019-nCoV), December 2019 to January 2020. Eurosurveillance. 2020. doi:10.2807/1560-7917.ES.2020.25.4.2000058

19. Souza WM de, Fletcher Buss L, Candido D da S, Carrera J-P, Li S, Zarebski AE, et al. Epidemiological and clinical characteristics of the early phase of the COVID-19 epidemic in Brazil. medRxiv - Imp Coll London. 2020. doi:10.1101/2020.04.25.20077396

20. USP. Epidemic simulator free app from São Paulo University. [cited 1 Jul 2020]. Available: https://ciis.fmrp.usp.br/covid19/epcalc/public/index.html

21. Ajzenman N, Cavalcanti T, Da Mata D. More Than Words: Leaders’ Speech and Risky Behavior during a Pandemic. SSRN Electron J. 2020. doi:10.2139/ssrn.3582908

22. Johns Hopkins University. COVID-19 Map - Johns Hopkins Coronavirus Resource Center. In: Johns Hopkins Coronavirus Resource Center. 2020.

23. Brazilian Ministery of Health. Brazilian national government COVID-19 website. 2020 [cited 6 Aug 2020]. Available: https://covid.saude.gov.br/

24. Ganem F, Mendes FM, Oliveira SB, Porto VBG, Araujo W, Nakaya H, et al. The impact of early social distancing at COVID-19 Outbreak in the largest Metropolitan Area of Brazil. medRxiv. 2020; 2020.04.06.20055103. doi:10.1101/2020.04.06.20055103

25. Canabarro A, Tenorio E, Martins R, Martins L, Brito S, Chaves R. Data-Driven Study of the COVID-19 Pandemic via Age-Structured Modelling and Prediction of the Health System Failure in Brazil amid Diverse Intervention Strategies. medRxiv. 2020; 2020.04.03.20052498. doi:10.1101/2020.04.03.20052498

26. Hou J, Hong J, Ji B, Dong B, Chen Y, Ward MP, et al. Changing transmission dynamics of COVID-19 in China: a nationwide population-based piecewise mathematical modelling study. medRxiv. 2020; 2020.03.27.20045757. doi:10.1101/2020.03.27.20045757

27. Yang Q, Yi C, Vajdi A, Cohnstaedt LW, Wu H, Guo X, et al. Short-term forecasts and long-term mitigation evaluations for the COVID-19 epidemic in Hubei Province, China. medRxiv. 2020. doi:10.1101/2020.03.27.20045625

28. Nussbaumer-Streit B, Mayr V, Dobrescu AI, Chapman A, Persad E, Klerings I, et al. Quarantine alone or in combination with other public health measures to control COVID-19: a rapid review. Cochrane database Syst Rev. 2020. doi:10.1002/14651858.CD013574

29. Richardson S, Hirsch JS, Narasimhan M, Crawford JM, McGinn T, Davidson KW, et al. Presenting Characteristics, Comorbidities, and Outcomes among 5700 Patients Hospitalized with COVID-19 in the New York City Area. JAMA - J Am Med Assoc. 2020. doi:10.1001/jama.2020.6775

30. Caramelo F, Ferreira N, Oliveiros B. Estimation of risk factors for COVID-19 mortality-preliminary results. medRxiv. 2020; 2020.02.24.20027268. doi:10.1101/2020.02.24.20027268

31. Bialek S, Boundy E, Bowen V, Chow N, Cohn A, Dowling N, et al. Severe Outcomes Among Patients with Coronavirus Disease 2019 (COVID-19) — United States, February 12–March 16, 2020. MMWR Morb Mortal Wkly Rep. 2020. doi:10.15585/mmwr.mm6912e2

32. Grasselli G, Zangrillo A, Zanella A, Antonelli M, Cabrini L, Castelli A, et al. Baseline Characteristics and Outcomes of 1591 Patients Infected with SARS-CoV-2 Admitted to ICUs of the Lombardy Region, Italy. JAMA - J Am Med Assoc. 2020. doi:10.1001/jama.2020.5394

33. Onder G, Rezza G, Brusaferro S. Case-Fatality Rate and Characteristics of Patients Dying in Relation to COVID-19 in Italy. JAMA-Journal of the American Medical Association. 2020. doi:10.1001/jama.2020.4683

34. Heck TG, Ludwig MS, Frizzo MN, Rasia-Filho AA, Homem de Bittencourt Jr PI. Suppressed anti-inflammatory heat shock response in high-risk COVID-19 patients: lessons from basic research (inclusive bats), light on conceivable therapies. Clin Sci. 2020;134: 1991–2017. doi:10.1042/CS20200596

35. Mehta P, McAuley DF, Brown M, Sanchez E, Tattersall RS, Manson JJ. COVID-19: consider cytokine storm syndromes and immunosuppression. The Lancet. 2020. doi:10.1016/S0140-6736(20)30628-0

36. Vaninov N. In the eye of the COVID-19 cytokine storm. Nat Rev Immunol. 2020. doi:10.1038/s41577-020-0305-6

37. Kissler SM, Tedijanto C, Lipsitch M, Grad Y. Social distancing strategies for curbing the COVID-19 epidemic. medRxiv. 2020; 2020.03.22.20041079. doi:10.1101/2020.03.22.20041079

